# Function is more reliable than quantity to follow up the humoral response to the Receptor Binding Domain of SARS-CoV-2 Spike protein after natural infection or COVID-19 vaccination

**DOI:** 10.1101/2021.06.02.21257975

**Authors:** Carlos A. Sariol, Petraleigh Pantoja, Crisanta Serrano-Collazo, Tiffany Rosa-Arocho, Albersy Armina, Lorna Cruz, E. Taylor Stone, Teresa Arana, Consuelo Climent, Gerardo Latoni, Dianne Atehortua, Christina Pabon-Carrero, Amelia K. Pinto, James D. Brien, Ana M. Espino

## Abstract

Both the SARS-CoV-2 pandemic and emergence of variants of concern have highlighted the need for functional antibody assays to monitor the humoral response over time. Antibodies directed against the spike (S) protein of SARS-CoV-2 are an important component of the neutralizing antibody response. In this work, we report that in a subset of patients—despite a decline in total S-specific antibodies—neutralizing antibody titers remain at a similar level for an average of 98 days in longitudinal sampling of a cohort of 59 Hispanic/Latino patients exposed to SARS-CoV-2. We also report that serum neutralization capacity correlates with IgG titers, wherein IgG1 was the predominant isotype (62.71%), followed by IgG4 (15.25%), IgG3 (13.56%), and IgG2 (8.47%) at the earliest tested timepoint. IgA titers were detectable in just 28.81% of subjects, and only 62.71% of subjects had detectable IgM in the first sample despite confirmation of infection by a molecular diagnostic assay. Our data suggests that 100% of seroconverting patients make detectable neutralizing antibody responses which can be quantified by a surrogate viral neutralization test. Examination of sera from 10 out of the 59 subjects which had received an initial first dose of mRNA-based vaccination revealed that both IgG titers and neutralizing activity of sera were higher after vaccination compared to a cohort of 21 SARS-CoV-2 naïve subjects. One dose was sufficient for induction of neutralizing antibody, but two doses were necessary to reach 100% surrogate virus neutralization in subjects irrespective of previous SARS-CoV-2 natural infection status. Like the pattern seen after natural infection, after the second vaccine dose, the total anti-S antibodies titers declined, however, neutralizing activity remained relatively constant for more than 80 days after the first vaccine dose. The decline in anti-S antibody titer, however, was significantly less in pre-exposed individuals, highlighting the potential for natural infection to prime a more robust immune response to the vaccine. Furthermore, our data indicates that—compared with mRNA vaccination—natural infection induces a more robust humoral immune response in unexposed subjects. However, this difference was significant only when neutralizing antibody titers were compared among the two groups. No differences were observed between naturally infected and vaccinated individuals when total anti-S antibodies and IgG titers were measured. This work is an important contribution to understanding the natural immune response to the novel coronavirus in a population severely impacted by SARS-CoV-2. Furthermore, by comparing the dynamics of the immune response after the natural infection vs. the vaccination, these findings suggest that a functional neutralizing antibody tests are more relevant indicators than the presence or absence of binding antibodies. In this context, our results also support standardizing methods of assessing the humoral response to SARS-CoV-2 when determining vaccine efficacy and describing the immune correlates of protection for SARS-CoV-2.

## Introduction

The COVID-19 pandemic presents an unprecedented challenge to the scientific community. At the same time, it is adding advancing our collective knowledge in molecular biology, epidemiology, and immunology at an accelerated speed. One of the crucial questions still under scrutiny is the magnitude and durability of the immune response to natural infection with SARS-CoV-2, especially given the fact that virus-specific antibody (ab) responses are relatively short-lived following SARS-CoV and common cold coronavirus infections (CCC) (Sette and Crotty 2020). Further complicating this scenario is the recent availability of new vaccine formulations, which are accessible to both previously infected and immunologically naïve individuals. The kinetics of the humoral response in vaccinees, both with and without prior SARS-CoV-2 exposure, is an area of active research with many outstanding questions.

To begin to address these questions, we followed a cohort of 59 individuals (volunteers or convalescent plasma donors) at different time points following natural infection with SARS-CoV-2. In addition, we chose a set of 7 of those individuals plus 3 additional subjects (n = 10) which we then compared with 21 uninfected-vaccinated subjects (n = 21). Serum samples for both vaccinated groups were collected between 12 and 28 days after each of the two doses of mRNA vaccine and a third sample was collected between 19 and 83 days after the second dose. Because the limited period of SARS-CoV-2 circulation, studies on the quantity, quality and extent of long-term memory responses are still underway. Recent works on the durability of the humoral immune response after the natural infection with SARS-CoV-2 showed the presence of neutralizing antibodies for several months (Dan et al. 2021, Figueiredo-Campos et al. 2020, L’Huillier et al. 2021, Lau et al. 2021, Wajnberg et al. 2020) or the persistence of IgG responses over the first few months after infection, which is strongly correlated with neutralizing antibody titer (Iyer et al. 2020, L’Huillier, Meyer, Andrey, Arm-Vernez, Baggio, Didierlaurent, Eberhardt, Eckerle, Grasset-Salomon, Huttner, Posfay-Barbe, Royo, Pralong, Vuilleumier, Yerly, Siegrist and Kaiser 2021). Since the onset of the COVID-19 pandemic, functional neutralization assays using serum antibodies has been severely limited due to the requirement for a biosafety level 3 (BSL-3) facility to grow SARS-CoV-2. However, in a relatively short period of time, several surrogate neutralization assays have become available with an excellent performance profile when compared to the classical focus reduction neutralization test (FRNT) (Jeewandara et al. 2021, L’Huillier, Meyer, Andrey, Arm-Vernez, Baggio, Didierlaurent, Eberhardt, Eckerle, Grasset-Salomon, Huttner, Posfay-Barbe, Royo, Pralong, Vuilleumier, Yerly, Siegrist and Kaiser 2021, Salazar et al. 2020, Schmidt et al. 2020, Tan et al. 2020, Taylor et al. 2021). For these studies, we choose the cPass SARS-CoV-2 Surrogate Virus Neutralization Test Kit (GenScript, USA) which measures the interaction of purified SARS CoV-2 spike protein receptor binding domain (RBD) with the extracellular domain of the human ACE2 receptor (Taylor, Hurst, Charlton, Bailey, Kanji, McCarthy, Morrison, Huey, Annen, DomBourian and Knight 2021). In our hands, this assay showed the best sensitivity and the lower false negative rate compared to five other assays (Tan, Saw, Chew, Huak, Khoo, Pajarillaga, Wang, Tambyah, Ong, Jureen and Sethi 2020). Furthermore, this assay was granted an Emergency Use Authorization (EUA) by the Federal Drug Administration (FDA) for the detection of SARS-CoV-2 neutralizing antibodies. Interestingly, we detected a small number of cases (n = 6) where neutralization activity was still present, although S-specific IgG titers were undetectable by our method (OD <.312).

Recently, debate has centered around the efficacy of the natural immune response to SARS-CoV-2 vs. mRNA vaccines. Our work which examines patients in a predominantly Latino population—confirms that following a natural infection neutralizing antibody titers remained detectable at high levels for 4 to 7 months. We also demonstrate that the quantity and the quality of the antibody response induced by the natural infection is significantly higher in titer of both binding and neutralizing antibodies when compared to the response induced by mRNA vaccination. There is limited information regarding the magnitude of the immune response to vaccines against SARS-CoV-2 in naive vs. pre-exposed subjects with clinical trial reports being limited in scope when addressing this issue (Baden et al. 2021, Sahin et al. 2020, Voysey et al. 2021, Walsh et al. 2020). Nevertheless, consistent with our findings presented here, a few reports suggest that antibody titers in previously infected persons trend or are significantly higher than in SARS-CoV-2 naïve persons (Bradley et al. 2021, Khoury et al. 2021, Krammer et al. 2021, Prendecki et al. 2021)., Together the results suggest that a single dose may be sufficient during the early stages of vaccine rollout in order to optimize vaccine availability worldwide.

## Material and Methods

### Cohorts

The samples in this study were derived from two main sources:

1-From adult volunteers (> 21 years old) participating in the IRB approved clinical protocol “Molecular Basis and Epidemiology of Viral infections circulating in Puerto Rico”, Pro0004333. Protocol was submitted to, and ethical approval was given by, Advarra IRB on April 21, 2020. This is a running 5 year protocol which encompasses the collection of blood samples from adults exposed or suspected to be exposed to viral infections. An Informed Consent Form and a study questionnaire also approved by the IRB were administered to the volunteers. From March 2020 to April 2021, we were able to follow up for serial samples with at least 59 subjects. From those 59, five (5) subjects received two doses of Pfizer’s vaccine and two received Moderna’s formulation. We also added three vaccinated subjects for a total of 10 (ID511, ID512 and ID297). From those three, two received Pfizer’s and one Moderna’s vaccine. All three subjects also consented to this study. In addition, a cohort of 21 vaccinated volunteers that were never exposed to SARS-CoV-2 were followed for 6 to eight months (Supplementary tables 1-3). Of these 21 vaccinated volunteers, eighteen (18) received Pfizer’s vaccine and three (3) received Moderna’s formulation. Those 21 subjects are part of the 59 subjects followed for months. During the follow-up period before vaccination, they never had symptoms or a positive serologic result. 2-De-identified blood samples received from local laboratories network and blood banks. These subjects were self-enrolled for the purpose of donating plasma for the treatment of COVID-19 patients. Subjects were verbally informed regarding the relevance of their participation in COVID-19 research, and were informed of the possibility that their deidentified samples may be used for research purposes. Subjects were given the opportunity to ask questions of blood bank workers regarding their participation. Furthermore, collected samples were handled using the standard blood donors’ protocols, and were accompanied by the blood bank’s signed consent form, which also detailed the possibility that samples would be used for research purposes. In addition, prior to receipt, samples were stripped of all identifiers so that the information cannot be traced back to the individual.

As expected, some of the exposed subjects had more symptoms than others, with fever and loss of smell and taste being the most common symptoms. However, in this cohort, subjects did not need hospitalization or additional medical support in an emergency room setting.

### Detection of SARS-CoV-2 IgM antibodies

CovIgM-Assay is an indirect ELISA for the determination of human IgM antibody class, which was optimized via checkerboard titration. This assay is a Laboratory Developed Test (LDT) with an Emergency Use Authorization (EUA) submitted to the U.S. Federal Drug Administration (FDA) (EUA202043). In summary, microplates were coated overnight at 4°C with 2μg/mL of recombinant SARS-CoV-2 S1-RBD protein (GenScript No. Z03483-1) in carbonate-bicarbonate buffer. Plates were washed 3 times with phosphate buffered saline (PBS) containing 0.05% Tween-20 (PBST) and blocked for 30 min at 37°C with 250μL/well of 3% Bovine Serum Albumin (BSA) in PBST. Diluted serum or plasma samples (1:100 in blocking buffer) were added in duplicates to the wells and incubated at 37°C for 30 min. The excess antibody was washed off with PBST. Horseradish peroxidase (HRP) labeled-mouse anti-human IgM-mu chain (Abcam) diluted 1:30,000 in PBST was added (100μL/well) and incubated for 30 min at 37°C. After another washing step, TMB solution was added (100μL/well) followed by 15 min incubation. The reaction was stopped by the addition of 50μL/well 10% HCl and the absorbance was measured at 450nm (A450) using a Multiskan FC reader (Thermo Fisher Scientific). In every CovIgM-Assay determination, four wells in which samples were replaced by 100μL/well of PBST were included as background control. Moreover, two in-house controls, a high positive control (HPC) and negative control (NC) were included. HPC and NC were prepared by diluting an IgM anti-SARS-CoV-2 at a concentration of 80μg/mL and 0.070μg/mL, respectively, in PBST containing 10% glycerol. The IgM anti-SARS-CoV-2 was purified from the plasma of a convalescent patient using 5/5 HiTrap IgM columns (GE Healthcare, USA). When the OD value of a serum or plasma sample at the working dilution (1:100) was equal or less than the cut-point (OD450= 0.229), the CovIgM-Assay in the sample was assumed to be negative.

### Detection of SARS-CoV-2 IgG antibodies

IgG antibodies were detected and quantified using the CovIgG-Assay (Espino et al. 2020). This assay is a Laboratory Developed Test (LDT) with an Emergency Use Authorization (EUA) submitted to the U.S. Federal Drug Administration (FDA) (EUA201115). It is an indirect ELISA for quantitative determination of human IgG antibody class, which was optimized by checkerboard titration. In summary, disposable high bind flat-bottomed polystyrene 96-wells microtiter plates (Costar, Corning MA No. 3361) were coated overnight at 4°C with 2μg/ml of recombinant SARS-CoV-2 S1-RBD/S2 protein (GenScript No. Z03483-1) in carbonate-bicarbonate buffer (Sigma Aldrich No. 08058). Plates were washed 3 times with (PBST) and blocked for 30 min at 37°C with 250μl/well of 3% non-fat, skim milk in PBST. Samples (serum or plasma) were diluted 1:100 in PBST; 100μL/well was added in duplicates and incubated at 37°C for 30 min. The excess antibody was washed off with PBST. Horseradish peroxidase (HRP) labeled-mouse anti-human IgG-Fc specific (GenScript No. A01854) diluted 1:10,000 in PBST was added (100μl/well) and incubated for 30 min at 37°C. After another washing step, a substrate solution (Sigma Aldrich No. P4809) was added (100μl/well) followed by 15 min incubation. The reaction was stopped with 50μl/well 10% HCl and the absorbance was measured at 492nm (A_492_) using a Multiskan FC reader (Thermo Fisher Scientific). In every CovIgG-Assay determination two in-house controls, a high positive control (HPC) and negative control (NC) were included. HPC and NC were prepared by diluting an IgG anti-SARS-CoV-2 at a concentration of 30μg/ml and 0.070μg/ml, respectively in PBST containing 10% glycerol. The IgG anti-SARS-CoV-2 was purified from plasma of a convalescent patient using a 5/5 HiTrap rProtein-A column (GE Healthcare, USA). When the OD value of a serum or plasma sample at the working dilution (1:100) was equal or less than the cutoff-point (OD492= 0.312), the CovIgG-Assay in the sample was assumed to be negative. However only samples with OD above of 0.499 were reported as having a titer within a range of 1:100 to ≥ 1:12,800.

For isotyping ELISAs, the conjugate was changed for the specific isotype as follows: anti-IgA (alpha chain specific-HRP (Sigma), anti-IgG1, 2, 3 and 4 Fc-specific-HRP (Southern Biotech). All conjugates were used in a 1:3,000 dilution.

### cPass SARS-CoV-2 neutralization antibody detection method

To determine the neutralizing activity of antibodies we used a surrogate viral neutralization test (C-Pass GenScript sVNT, Piscataway NJ) (Tan, Saw, Chew, Huak, Khoo, Pajarillaga, Wang, Tambyah, Ong, Jureen and Sethi 2020, Taylor, Hurst, Charlton, Bailey, Kanji, McCarthy, Morrison, Huey, Annen, DomBourian and Knight 2021). Briefly, serum or plasma samples were diluted according to manufacturer’s instructions and incubated with soluble SARS-CoV-2 receptor binding domain (RBD-HRP) antigen for 30 minutes, mimicking a neutralization reaction. Following incubation, samples were added to a 96 well plate coated with human ACE-2 protein. RBD-HRP complexed with antibodies are removed in a wash step. The reaction is developed with tetramethylbenzidine (TMB) followed by a stop solution allowing the visualization of bound RBD-HRP to the ACE2. Since this is an inhibition assay, color intensity is inversely proportional to the amount of neutralizing antibodies present in samples. Data is interpreted by calculating the percent of inhibition of RBD-HRP binding. Samples with neutralization activity of ≥30% indicates the presence of SARS CoV-2 RBD-interacting antibodies capable of blocking the RBD-ACE2 interaction thus inhibiting viral entry into host cells. While this assay measures the blocking activity of those antibodies, for consistency and clarity this activity is referred to throughout the text as ‘percentage of neutralization’.

### Statistical Methods

Statistical analyses were performed using GraphPad Prism 7.0 software (GraphPad Software, San Diego, CA, USA). The statistical significance between or within groups evaluated at different time points was determined using two-way analysis of variance (ANOVA) (Tukey’s, Sidak’s or Dunnett’s multiple comparisons test) or unpaired t-test to compare the means. The p values are expressed in relational terms with the alpha values. The significance threshold for all analyses was set at 0.05; p values less than 0.01 are expressed as P<0.01, while p values less than 0.001 are expressed as P<0.001. Similarly, values less than 0.005 are expressed as P<0.005. Cohen’s Kappa agreement follow Landis and Koch scale. The values (κ) were considered as follows: poor agreement, κ<0.02); fair agreement, κ=0.21 to 0.4; moderate agreement, κ=0.41 to 0.6; substantial agreement, κ=0.61 to 0.8; very good agreement, κ=0.81 to 1.0.

## Results

### Sample collection

Subjects were enrolled and samples were collected as participants became willing and available. However, the time between serial samples was very similar for all subjects. The average time between the time of the documented infection and the first samples (n=59) was 40.37 days (minimum 12 days, maximum 97 days and two extreme cases with 127 and 176 days for a median of 38 days). Once the subjects entered in the cohort, the average time between the first and the second samples (n=59) was 67.86 days (minimum 7 days, maximum 111 days, median 67.5 days). The average time between the second and the third samples (n=12) was 99.5 days (minimum 63 days, maximum 159 days, median 95 days) (Supplementary table S1). From the two subgroups, exposed-vaccinated and unexposed-vaccinated, serum samples were collected between 15 to 20 days after each dose. In addition, a third sample from all 21 unexposed and from 8 out of the 10 pre-exposed participants was collected between 19 and 83 days after the second dose (average of 40.1 and of 81.6 days for the unexposed and pre-exposed groups respectively) or an average of 60.3 and 100.5 days after the first vaccine dose for the unexposed and pre-exposed groups respectively (Supplementary table S2). Highly relevant for our findings is that the sample used as baseline in the pre-exposed before vaccination, was collected in average 142 days after the confirmed infection (minimum 67 days, maximum 310 days, median 126.4 days) (Supplementary table S3).

### SARS-CoV-2 specific IgG titers decline over time

Overall, the IgG titers in the cohort of 59 subjects were significantly higher (geometric mean 1072) in the first set of samples than the second set of samples (geometric mean 618) (p< 0.0473) or the third set of samples (geometric mean 537) (p< 0.0474). We observed no significant differences between titers measured in the second and third sets of samples (p < 0.3085) (Figure 1A). The results are reported as OD450 in supplementary figure 1A and agree with estimated titers (Supplementary table S4).

**Figure 1:**
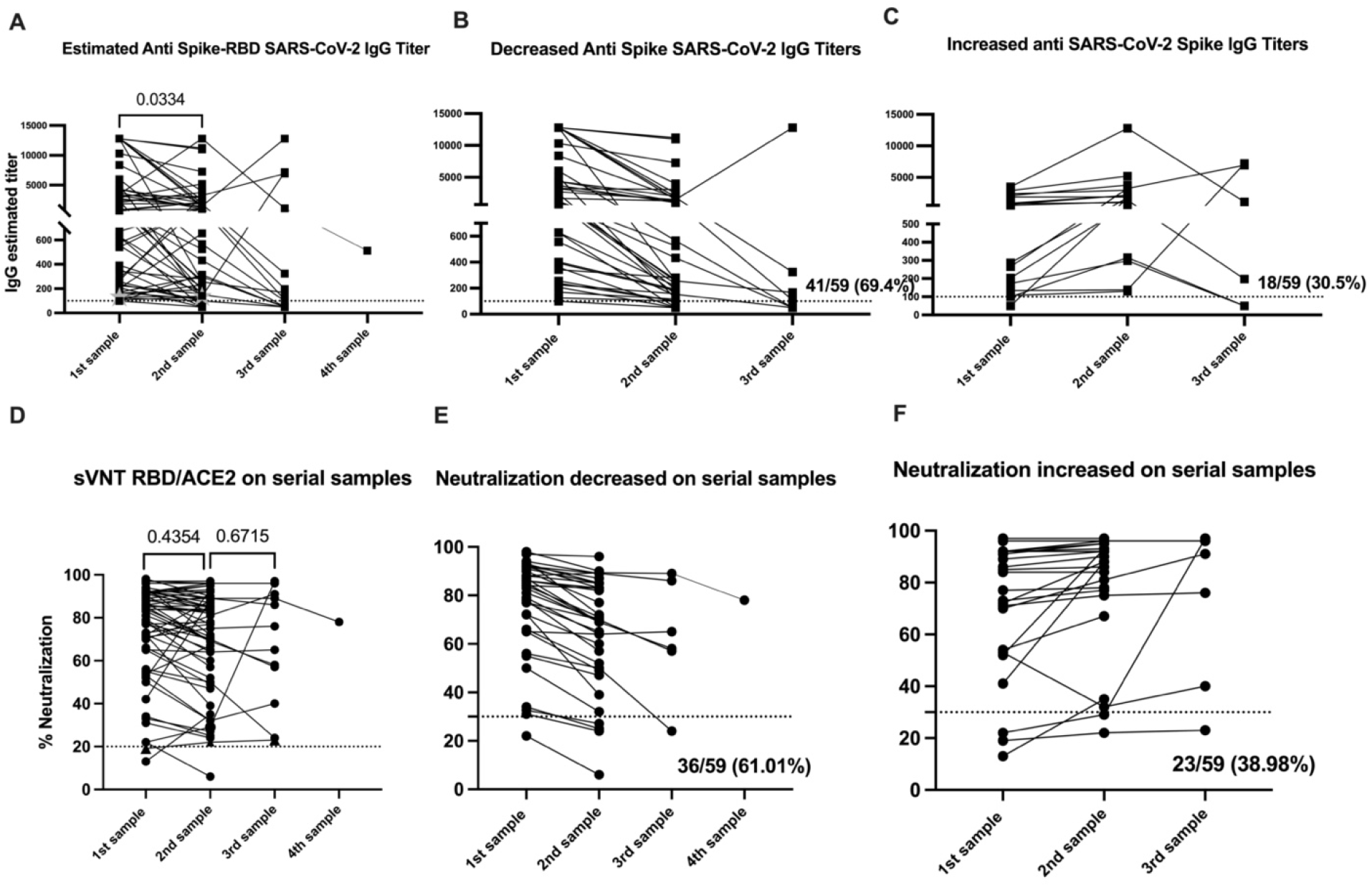
SARS-CoV-2 specific antibody titers decline over time, while neutralization ability is retained. The threshold for the total antibodies was 0.312. The threshold for IgG titers was 1:100 and for the blocking activity was 30%. Statistical significance was determined by 2way ANOVA multiple comparisons was used to test for increase or decrease among samples. P<0.05 was considered significant. Samples 1 and 2 include the 59 subjects in the initial cohort before vaccination. Sample 3 encompass the 15 subjects from whom a collection of a third sample was completed.

Of the 59 subjects naturally exposed to the virus, 40 (67.8%) experienced a decrease in IgG titers (Figure 1B) while 19 (32.2%) showed an increase in the IgG titers from the first to the second set of samples (Figure 1C). We found no relationship between the elapsed time from initial diagnosis to first sample collection and the change (e.g. increase or decrease) in IgG titers between sample collections (Supplementary figure 2). From these results, we concluded that the differences in the IgG titers in those groups from the first to the second set of samples were not attributable to the time between collection. We also found no relationship between the elapsed time between the first and second sample collection for both groups (Supplementary figure 2). We identified three subjects (ID137, ID195, and ID367) showing a unique trend towards an increase in IgG titers of 2.13-, 8.65-, and 52.1-fold, respectively, between the second and third sampling collections. Particularly, volunteer ID195 exhibited an initial 8.65-fold decrease in IgG titer between the first and second sample collection (50 days elapsed) followed by an increase in IgG titer between the second and third sample collection (75 days elapsed).

### SARS-CoV-2 specific IgM titers decline over time

Among the 59 subjects in our cohort, 37 (62.71%) had detectable IgM titers in the first set of samples, while 18 (30.50%) had detectable IgM titers in the second set of samples (Supplementary figure 1H). In five subjects out of the 12 where a third sample was collected, IgM titers were still detectable. In some cases, subjects developed an IgM response for first time (in volunteer ID313 IgM was detected as early as 12 days after the presumptive diagnosis and persisted up to 192 days, or roughly 6.4 months). Overall, IgM titers showed a consistent pattern of decline in the second sample for most individuals (86.44%). Only one subject (ID265) showed no appreciable change in IgM titers between the first and second sample collection (68 days elapsed). One subject (ID313) displayed no measurable IgM titer at the time of the first and second sampling (106 days elapsed), but appeared positive for IgM titers in the third sample (146 days elapsed). We also found that in the second set of samples, 3 subjects out of 59 (5.08%) displayed detectable IgM titers which were absent detectable IgG titers. Subject ID312 showed detectable IgM titers, but borderline IgG titers results in the third sample collected 57 and 69 days after the first and second dose, respectively (86 and 98 days after the presumptive diagnosis). Subject ID105 still had detectable IgM titers 192 days after the presumptive diagnosis was made. The earliest time point with detectable IgM titers was 12 days after the presumptive infection (ID166), followed by 13 days (ID180) and 14 days (ID179) after diagnosis. In general, IgM was detected in 37 subjects (77.97%) in the first set of samples (43 days post presumptive infection). In 18 subjects (57.63%), IgM was detected in the first and second set of samples (104 days post presumptive infection). In 4 subjects (6.77%) no IgM was detected in any of the serial samples collected.

### IgG titers—but not IgM or IgA titers—correlate with neutralizing activity

As described previously, the correlation between estimated IgG titers by the CovIgG-Assay and the neutralization capacity as measured by the Focus Reduction Neutralization Test (FRNT) is extremely strong (Espino, Pantoja and Sariol 2020). For this work, we performed same analysis examining the correlation between IgG titers and functional neutralization capacity, obtained in these studies using the surrogate assay cPass SARS-CoV-2 neutralization antibody detection method. By applying a Kappa analysis, we first aimed to determine if both techniques agree when classifying positive and negative samples using <100% and >30% as cutoff for the IgG titers and percentage of neutralization respectively. We found moderate agreement between IgG titer and neutralization capacity, with a Cohen’s Kappa value of 0.4304 (Supplementary figure 3A). We then aimed to determine whether both techniques agree when classifying samples with high IgG titers and high neutralizing antibody titer. Similarly, we found moderate agreement between IgG titer and neutralization capacity, with a Cohen’s Kappa value of 0.5402 (Supplementary figure 3B). We completed the same analysis for IgM and IgA titers to explore the contribution of those antibody subclasses to total neutralization capacity. We found that both techniques (IgM titer and cPass) have a fair agreement when classifying positive and negative samples (Cohen’s Kappa = 0.2391), while the IgA titer and the neutralization assay showed only a slight agreement (Cohen’s Kappa = 0.0618) (Supplementary figure 3C-D).

### Neutralizing activity remains constant over time

To determine the durability of the neutralizing antibody response, we examined the neutralization capacity in our longitudinally collected samples. Our results showed consistent neutralizing antibody titers over time, with no change in the neutralization potential from the first (geometric mean 68.08%) to the second (geometric mean 63.89%) sample. Similarly, we saw no appreciable decline in neutralization potential from the second (geometric mean 63.89%) to the third (geometric mean 60.36%) sample (Figure 1D and supplementary table S4). We did, however, identify two distinct trends in the kinetics of serum neutralization potential over time. Similar to our findings with total IgG titers, in the first collected sample we found a decrease in the neutralizing activity relative to the second sample in 61.01% (36 out of 59) of the subjects. Conversely, 38.98% of subjects (23 out of 59) showed a decrease in neutralization activity (Figures 1E and F) during the same timeframe. While the percentage of subjects experiencing an increase or a decrease in neutralization capacity and IgG titers between samples was similar, the change in neutralization capacity was less pronounced and not significant compared with significant changes in the IgG titers (Figure 1B). From these findings, we concluded that the neutralizing capacity remains relatively constant during the time we followed this cohort.

Similarly, we compared the neutralization potential of sera from subjects in the second and the third samples for the few subjects (n = 3) for which we were able to obtain a third sample. We identified one subject (ID313) showing a different pattern, with a 3.34-fold (68%) increase in neutralizing activity from the second to the third sample. Another two subjects showed an increase in IgG titers, but displayed a very limited increase in neutralizing activity of 1.2-fold (ID135) and 0-fold (ID195). Despite the variability in IgG titers, neutralizing activity remained over 50% in a majority (90%) of all three samples. The distinctive serological and neutralization pattern for subject ID313 appears to be strongly related to the clinical evolution (Supplementary figure 3).

We also identified 11 subjects without detectable SARS-CoV-2 specific IgG titers which showed some degree of neutralization ranging from 36% to 76%. Six out of those 11 subjects had no detectable total IgG. On the other hand, there were 3 subjects with detectable IgG titers capable of binding SARS-CoV-2 S protein, but with very limited or absent neutralization capacity (Supplementary table S4).

### Natural infection induces high quality antibodies than one vaccine dose

Next, we wanted to compare the magnitude of the humoral immune response to naturally acquired SARS-CoV-2 infection to the mRNA-based COVID-19 vaccinations in unexposed subjects. For this purpose, we choose samples from 25 participants out of the 59 with the first sample collected between 12 and 39 days after the confirmed infection with SARS-CoV-2 (average 26.23 days) and from 21 unexposed participants that received two doses of the Pfizer-BioNTech vaccine. Samples for the unexposed subjects were collected an average of 17.1 and 14.1 days after the first and the second dose, respectively. As shown in Figure 2A, the mean time elapsed between the first sample collection after infection was significantly higher than the time elapsed between the first sample collected after the vaccination in the unexposed cohort (*p<*0.0001). Despite this delay, we found that the total anti-S antibodies and the total IgG titers were comparable after the infection or the first vaccine dose in the unexposed participants (Figures 2B and D). However, the quality of the antibodies measured by the surrogate neutralization assay showed a neutralizing activity significantly higher in the naturally infected group compared with the unexposed-vaccinated group (p<0.0003). This indicated to us a better quality of the antibodies induced by naturally acquired infection when compared to vaccine-induced neutralizing antibody activity (Figure 2D). As showed in Figures 2B and 2C, two vaccine doses in unexposed individuals were necessary to significantly increase the total antibody titers and IgG titers compared to individuals in the pre-exposed group (p<0.0004). The magnitude of neutralization was also significantly increased in pre-exposed individuals, but more modestly than the quantity (p<0.0294), suggesting that the increase in antibody quantity induced by the two vaccine doses was not accompanied by a similar increase in the quality of the neutralizing antibody response (Figure 2D).

**Figure 2:**
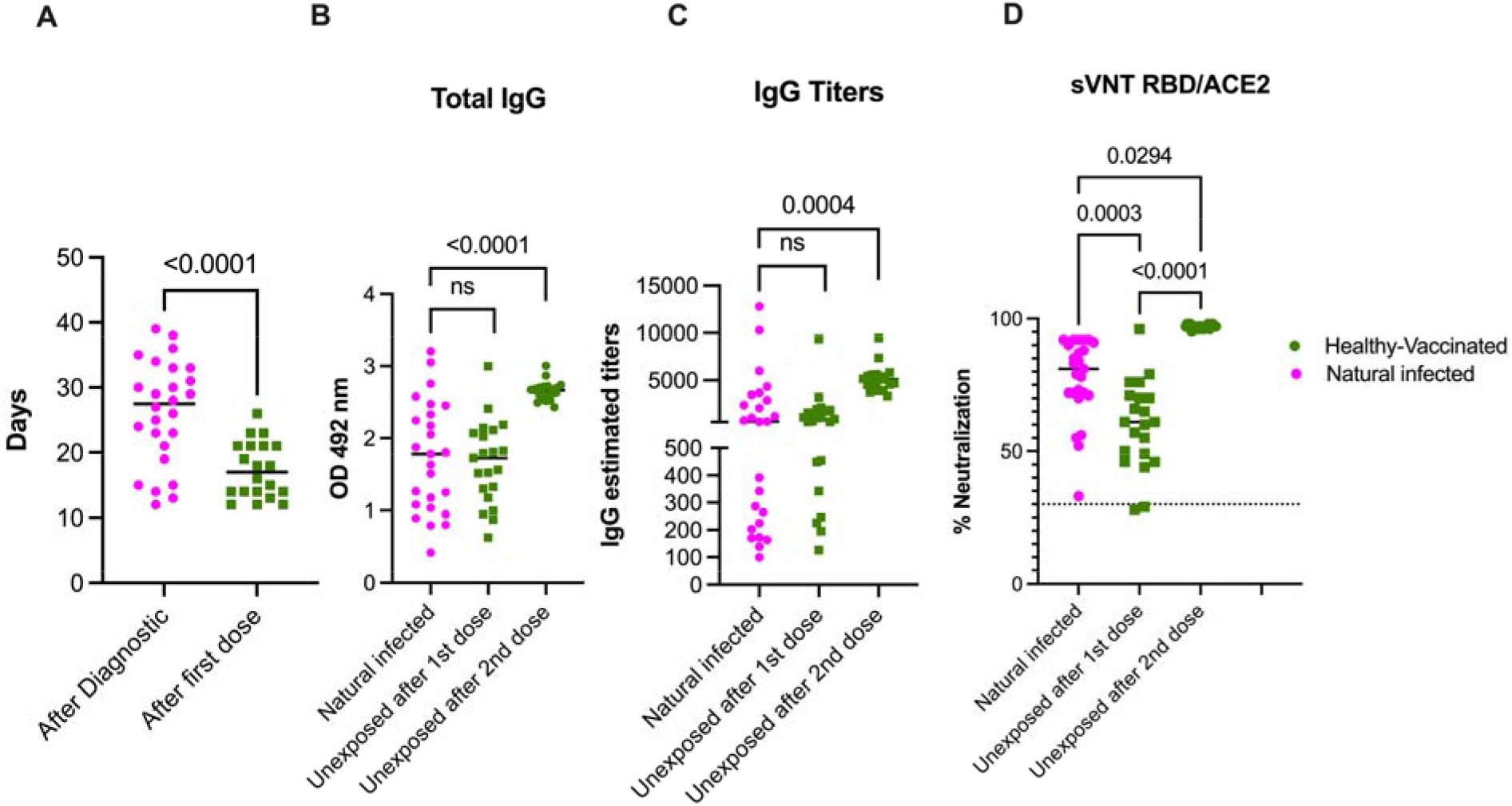
Naturally acquired SARS-CoV-2 infection primes an immune response superior to a single COVID-19 vaccine dose. Panel A shows the mean time of sample collection following natural infection (n=25) or after the first vaccine dose (n=20). In panel B and C, results from the total anti-Spike protein and the IgG titer measured by Enzyme-linked immunosorbent assay and expressed as OD or titers respectively are presented. The threshold for the total antibodies was 0.312 and the threshold for IgG titers was 1:100. All participants, except one, with previous exposure to SARS-CoV-2 showed detectable antibodies and measurable titers at baseline. Because the threshold 1:100 of our titration assay, the IgG titers at baseline in the unexposed subjects—which had no detectable S-specific antibodies—were set arbitrarily to 50. Panel D shows the blocking activity of serum antibodies expressed as percentage of neutralization by using a surrogate viral neutralization test (sVNT). The cutoff for this assay was 30%. As is shown, only one sample in the pre-exposed group contained antibodies below the threshold reported as more than 30% of neutralization. Also, while the distribution of antibodies and titers covers the full Y axis, values in both panel B and C, and in panel D same samples are grouped on the top values area. Two-way ANOVA multiple comparisons or unpaired T test analysis was used to test for increases or decreases among samples. P<0.05 was considered significant. Twenty-five participants (Natural infected) out of the 59 with the first sample collected between 12 and 39 days after the confirmed infection with SARS-CoV-2 were selected for comparison with the 21 unexposed-vaccinated subgroup (Healthy-vaccinated).

### Neutralization is sustained in naïve and pre-exposed-vaccinated subjects

Samples were collected between 12 to 28 days after each dose with a mean of 19 days and of 14 days for the pre-exposed group and of 12 days and 26 days for the unexposed groups after the first and second dose respectively. An additional third sample from all 21 unexposed individuals and from 8 out of the 10 pre-exposed individuals was collected between 19 and 83 days after the second dose, respectively (Supplementary table S2). For the first sample collected following the first dose, there were no significant differences in the time elapsed between sample collections for the pre-exposed and unexposed subjects. However, there was a significant difference (*p<*0.0001) in the time elapsed between sample collections following the second dose (third sample) between the pre-exposed and unexposed groups (Supplementary figure S4). The geometric mean baseline IgG titers in the pre-exposed population was 726 (range: 125 to 7191) and increased to a geometric mean of 5239 (range: 3408 to 6586) after the first dose (Figure 3B and supplementary tables S5 and S6). After the second dose, the geometric mean decreased to 3980 (range: 2273 to 5847), and we observed no significant difference in IgG titers after the first dose. On the other hand, the 21 vaccinated, unexposed subjects were negative for S-specific IgG at baseline. After the first dose, the IgG titers significantly increased to a geometric mean of 832 (range: 196 to 9365, p<0.0001) and after the second dose, those values significantly increased (p<0.0001) to a geometric mean of 5446 (range: 3346 to 10,239) (Figure 3B).

**Figure 3:**
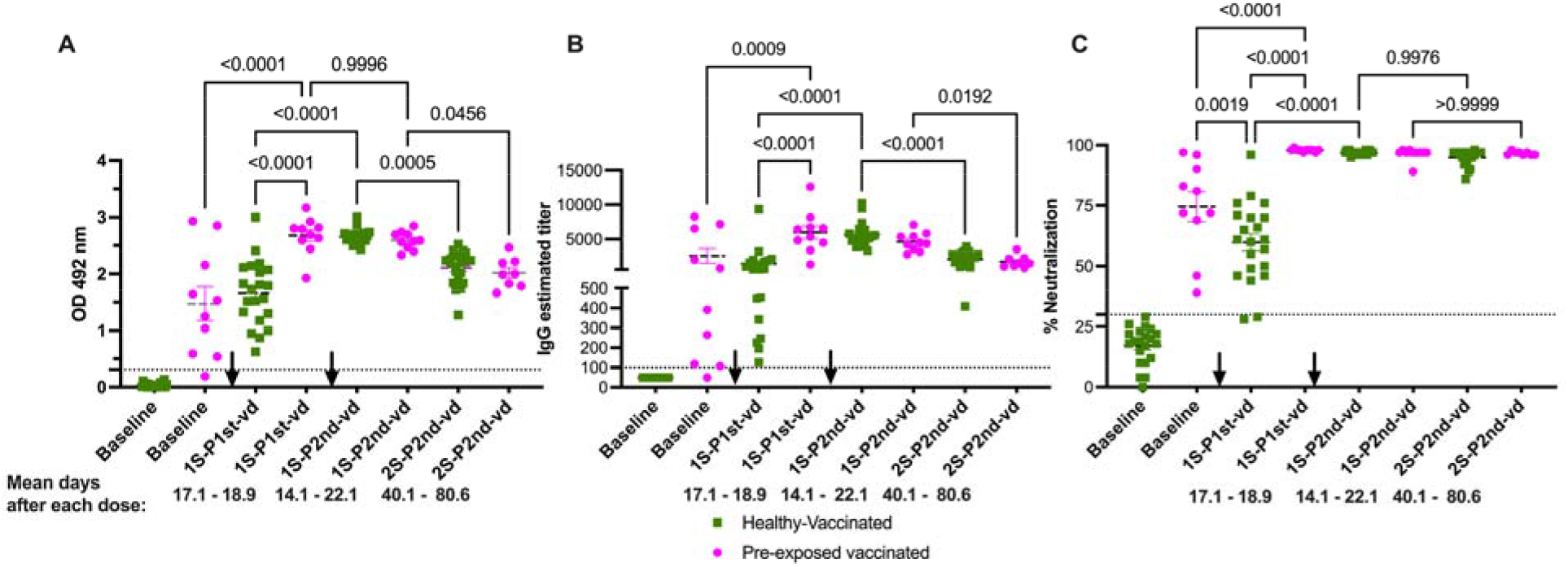
Functional neutralization assays are better predictors of the humoral immune response to COVID-19 mRNA-based vaccinations. Samples are described as 1st or 2nd samples after 1^st^ or 2^nd^ vaccine dose (1S-P1st-vd, 1S-P2nd-vd or 2S-P2nd-vd) and the mean time of samples collection is shown. Panels A and B show the total antibody and IgG titers, respectively, after full vaccination with two vaccine doses. Antibody levels and titers significantly decay in both groups in a second sample collected after the second vaccine (average of 60.3 and 100.5 days after the first vaccine dose for the unexposed and pre-exposed groups respectively). Despite the difference in sampling time between the two groups, there were no significant differences in the levels of antibodies or titers between groups in the 2S-P2nd-vd. Panel C shows antibody blocking capabilities measured by a surrogate viral neutralization assay (sVNT). Highly relevant is the finding that the blocking baseline activity of the pre-exposed individuals is significantly higher than the basely blocking activity induced by the first vaccine dose in unexposed individuals. In addition, two vaccine doses were necessary in the unexposed cohort to induce same percentage of neutralization achieved by just the first dose in the pre-exposed group. The magnitude of neutralization remained at similar levels until the last time point evaluated in both groups, confirming that the surrogate neutralization test is more suitable to determine the efficacy of the humoral immune response to the vaccine. The threshold for the total antibodies was 0.312. The threshold for IgG titers was 1:100 and for the blocking activity was 30%. Statistical significance was determined by two-way ANOVA multiple comparisons to test for increase or decrease among samples. p<0.05 was considered significant. The black arrows indicate the moment of vaccine administration related to the timing of sample collection. Healthy-vaccinated (n=21) Pre-exposed vaccinated (n=10).

In the second sample, which was collected after the second dose (third sample) in the unexposed group, the geometric mean of the titers was 1518 (range: 409 to 3278). In the pre-exposed group, the geometric mean of the titers was 1323 (range: 568 to 3536). In both groups, we observed a a significant decrease from the IgG titers detected in the first samples relative to titers after the second dose (*p<*0.0001 and *p*=0.0192 for the unexposed and pre-exposed groups, respectively).

In our cohort, the total IgG values were consistent with reported IgG titers (Figure 3A). We looked first at the IgG1 isotype, the main contributor to the total IgG in the cohort of 59 individuals. The first dose induced a significant increase in this isotype for both groups (p<0.0018 and p<0.0001 for the unexposed and pre-exposed vaccinated groups, respectively). However, the effect of the boost was significantly higher in the pre-exposed group (p<0.0001) suggesting a role for natural infection in this significant difference. Remarkably, the second dose appeared to provide a benefit in boosting IgG1 titers in the unexposed, vaccinated group only (p<0.0001). IgG1 values after the second dose in the unexposed, vaccinated group reached values comparable to that of the pre-exposed vaccinated group after just one dose. We observed no significant differences in the levels of IgG1 between groups following the second dose (Supplementary Figure 5).

The geometric mean baseline of neutralization activity in the pre-exposed population was 69.46% (range: 39 to 97%) and increased significantly (p<0.0001) to a geometric mean of 97.99% (range: 97 to 98%) after the first dose (Figure 3C and Supplementary Table S5). However, following the second dose, the values remained similar in range, with a mean of 97.19%. On the other hand, the 21 naïve-vaccinated persons were negative for neutralization at baseline (geometric mean: 15%). After the first dose, neutralization significantly increased (p<0.0001) to a geometric mean of 57.34% (range: 28% to 76%, with one outlier of 96%). The second dose produced an additional significant boost (p<0.0001) to a geometric mean of 96.85% (in a range from 95% to 98%) (Figure 3C). Contrary to the trend we observed in total antibody titers and IgG titers (Figures 3A and B), the neutralizing activity was retained at very similar level in both groups in the third sample collected. The geometric mean for the unexposed group was 94.5% (in a range from 86% to 98%), while the pre-exposed group had a geometric mean of 96.62% (in a range from 96% to 98%). Though there was no significant difference in neutralization capacity between groups, nine (9) subjects in the unexposed group showed values lower than 5% neutralization. This resulted in a 1.02-fold decrease in the value of neutralization capacity in the unexposed group, while there were no changes in neutralization capacity the pre-exposed cohort.

Among the previously exposed subjects we examined, 5 out of 10 (50%) retained detectable IgM at baseline (i.e. the time of the first sampling). IgM titer did not appear to be boosted by the first vaccine dose, and titers decreased after the second dose. On the other hand, the first dose did appear to induce a significant increase (p<0.0001) in the IgM values in the unexposed subjects. Those values were boosted only in two subjects, but as expected, were not modified in any of the other 19 subjects (Supplementary Figure 5). Eight (8) out of the 21 unexposed patients (38.09%) had no detectable IgM after the first dose. Only one patient failed to develop measurable IgM antibodies after the two vaccine doses.

Finally, we looked at the contribution of the IgA isotype to the immune response after vaccination. Interestingly, we found that this isotype was significantly boosted in both groups, pre-exposed (p<0.0187) and unexposed groups (p<0.0010) after the first vaccine dose. In addition, the increase in IgA titers was significantly higher in the pre-exposed (p<0.0176) vaccinated group compared to the unexposed, vaccinated group. The second boost resulted in an additional significant increase in IgA titers in the unexposed, vaccinated population but not in the pre-exposed vaccinated group (Supplementary Figure 5).

## Discussion

Our study followed a cohort of 59 subjects with prior exposure to SARS-CoV-2 with the goal of describing the kinetics of the humoral immune response to natural infection over time. This study uniquely examined a population of Hispanic/Latino persons disproportionately impacted by the COVID-19 pandemic. We compared the kinetics of this antibody response in the context of individuals with naturally acquired infection (pre-exposed) and unexposed individuals following vaccination. None of the exposed subjects in our cohorts required hospitalization and only had mild to moderate symptoms. Because of that, we found no differences in the serological response according to symptoms severity. Consistent with other reports, we found that antibody titers tended to wane over time and added to a growing body of evidence suggesting that functional neutralization assays should serve as the gold standard for evaluating vaccine efficacy in lieu of antibody binding quantification. Furthermore, we found that pre-exposed individuals were able to mount an antibody response after just one vaccination dose that was equivalent to a two-vaccine dose regiment in unexposed individuals. These findings have important implications for defining the correlates of protection for SARS-CoV-2, as well as recommendations for future public health guidelines and vaccine distribution efforts on a global scale.

One limitation of our work is the limited number of subjects sampled following natural infection or vaccination. However, we were able to draw statistically significant conclusions from our studies using 59 individuals. Additionally, our findings in this limited dataset are consistent with previous reports, which have made great contributions to our understanding of the immunological response to SARS-CoV-2 with a similar number of subjects (Bradley, Grundberg, Selvarangan, LeMaster, Fraley, Banerjee, Belden, Louiselle, Nolte, Biswell, Pastinen, Myers and Schuster 2021, Geers et al. 2021, Krammer, Srivastava, Alshammary, Amoako, Awawda, Beach, Bermúdez-González, Bielak, Carreño, Chernet, Eaker, Ferreri, Floda, Gleason, Hamburger, Jiang, Kleiner, Jurczyszak, Matthews, Mendez, Nabeel, Mulder, Raskin, Russo, Salimbangon, Saksena, Shin, Singh, Sominsky, Stadlbauer, Wajnberg and Simon 2021, Prendecki, Clarke, Brown, Cox, Gleeson, Guckian, Randell, Pria, Lightstone, Xu, Barclay, McAdoo, Kelleher and Willicombe 2021).

We also acknowledge that setting up a longitudinal cohort study is always a challenge. Particularly for COVID-19, it imposed additional difficulties due to the lockdowns, social distancing measures, stigma associated with positive testing, and other significant barriers. However, we assert that the limitations regarding the sampling sequence do not detract from the significance of our findings.

Notably, our results contrast with reports describing a short persistence of neutralizing antibodies in plasma donors (Annen et al. 2021), but are in agreement with recent work indicating that neutralizing antibodies may persist longer (Dan, Mateus, Kato, Hastie, Yu, Faliti, Grifoni, Ramirez, Haupt, Frazier, Nakao, Rayaprolu, Rawlings, Peters, Krammer, Simon, Saphire, Smith, Weiskopf, Sette and Crotty 2021, Klingler et al. 2020, Wajnberg, Amanat, Firpo, Altman, Bailey, Mansour, McMahon, Meade, Mendu, Muellers, Stadlbauer, Stone, Strohmeier, Aberg, Reich, Krammer and Cordon-Cardo 2020). Another work showed a long-term stabilization of anti-Spike IgG value and nAbs lower than in early days post symptoms onset in a hospitalized cohort (Dispinseri et al. 2021). The effect we are seeing in the samples with a decrease in the total antibodies and titers in the second sample may be also a stabilization at a plateau. We have followed up samples from 8 out of the 10 pre-exposed vaccinated subjects, but unfortunately, alterations in the humoral response due to vaccination of these subjects limit our interpretation of these results. Interestingly, the same group reported that nAbs are a correlate of survival and that nAbs and, that anti-spike IgG persists in the vast majority of recovered patients regardless of disease severity, age, and co-morbidities for up to eight months from symptoms onset (Dispinseri, Secchi, Pirillo, Tolazzi, Borghi, Brigatti, De Angelis, Baratella, Bazzigaluppi, Venturi, Sironi, Canitano, Marzinotto, Tresoldi, Ciceri, Piemonti, Negri, Cara, Lampasona and Scarlatti 2021). A longer follow up period would further our understanding of the antibody kinetics in a long-term period

We were able to show a similar trend in our cohort, with sustained neutralizing activity during the frame time of this study. The sustained neutralization capacity we observed remains highly relevant, despite the significant decline of IgG titers that we observed in this cohort. In addition, we found that some subjects with undetectable IgG (n=6) and IgG titers (n=11) retain measurable neutralization activity, ranging from 32 to 76 %, as measured by a surrogate virus neutralization assay. This finding is consistent with previous reports, suggesting that SARS-CoV-2 serological assays may be poorly-suited for prediction of serum neutralization potency, a metric necessary to facilitate the establishment of the appropriate serologic correlates of protection against SARS-CoV-2 (Muecksch et al. 2020). Our results suggest that functional assays measuring neutralization potential should be implemented in studies of vaccine efficacy at the population level.

From a technical point of view, the discrepancies between samples without detectable antibodies but with neutralizing capabilities may be explained by differences in assays’ sensitivity. In our case, we use the same source of recombinant proteins for the antibodies and surrogate neutralization assays. However, the serological assays include the full S1 and S2 regions of the Spike protein, which includes the RBD, to coat the plate. The neutralization assay, however includes only the S1/RBD in suspension. It has been well documented that the binding of the protein to the plate results in altered antigen accessibility with a consequent presentation of different antigenic sites compared to native proteins (de Thier et al. 2015, Güven et al. 2014, Mannik et al. 1997, Taylor, Hurst, Charlton, Bailey, Kanji, McCarthy, Morrison, Huey, Annen, DomBourian and Knight 2021). Nevertheless, we showed a 93.7% correlation between IgG titers and neutralization measured with a cPass SARS-CoV-2 Neutralizing Antibody Detection kit.

There are a limited number of publications on the contribution of different antibody isotypes to the immune response to this novel coronavirus. Early studies reported that spike-and RBD-specific IgM, IgG1, and IgA antibodies were detected in most subjects early after infection, with all samples displaying neutralizing activity and IgM and IgG1 contributing most to neutralization (Klingler, Weiss, Itri, Liu, Oguntuyo, Stevens, Ikegame, Hung, Enyindah-Asonye, Amanat, Baine, Arinsburg, Bandres, Kojic, Stoever, Jurczyszak, Bermudez-Gonzalez, Nádas, Liu, Lee, Zolla-Pazner and Hioe 2020). A recent work reported that in a hospitalized cohort early presence of anti-RBD anti-spike IgA positively correlated with reduced persistence of SARS-CoV-2 RNA in naso-pharyngeal swabs (Dispinseri, Secchi, Pirillo, Tolazzi, Borghi, Brigatti, De Angelis, Baratella, Bazzigaluppi, Venturi, Sironi, Canitano, Marzinotto, Tresoldi, Ciceri, Piemonti, Negri, Cara, Lampasona and Scarlatti 2021). Other work reported that early SARS-CoV-2-specific humoral responses were dominated by IgA antibodies and that virus-specific antibody responses included IgG, IgM, and IgA. Furthermore, some studies have found that the IgA isotype contributes to virus neutralization to a greater extent compared with IgG (Sterlin et al. 2021). In agreement with our results, recent work from India, a heavily impacted country by the pandemic found that RBD-specific IgG but not IgA or IgM titers, correlated with neutralizing antibody titers and RBD-specific memory B cell frequencies (Nayak et al. 2021).In our work, we found that IgG1 was the predominant isotype, while the IgA response was more limited. However, considering the non-significant changes in the IgA levels from the first to the second sample, a role for IgA in sustained neutralization activity cannot be ruled out. On the other hand, in the majority of subjects in this cohort, IgM showed an expected trend to decline in the second collected sample. Two out of four subjects (ID265 and ID382) which were IgG-/IgM+, also had detectable neutralizing activity with detectable IgM both two and four months after the first samples were collected. These cases suggest that in some individuals, IgM may contribute to sustained neutralization capacity, as has been described before (Klingler, Weiss, Itri, Liu, Oguntuyo, Stevens, Ikegame, Hung, Enyindah-Asonye, Amanat, Baine, Arinsburg, Bandres, Kojic, Stoever, Jurczyszak, Bermudez-Gonzalez, Nádas, Liu, Lee, Zolla-Pazner and Hioe 2020). This result also corresponds with a Kappa analysis suggesting a fair Cohen’s Kappa agreement between IgM titers and neutralization capacity. Additional isotype-specific depletion experiments are needed to determine the role of these antibodies in SARS-CoV-2 neutralization. Using previous experience from our group (Serrano-Collazo et al. 2020, Steffen et al. 2020) those experiments are underway using a larger number of well characterized individuals.

While the number of subjects in our vaccinated cohort (both unexposed and previously exposed subjects) is limited, we show that vaccination induces a higher boost in the magnitude of the humoral immune response, both at the level of S-specific IgG and neutralization ability in the pre-exposed individuals compared to the naïve group. Our findings also indicate that the second vaccine dose did not expand the S-specific antibodies, the total IgG titers, or the neutralization capacity of blocking antibodies beyond the peak reached after the first dose in the case of the pre-exposed cohort. One subject (ID112) received the Moderna formulation (ID112) was identified as unexposed and without any known exposure to the SARS-CoV-2, reach values in all three determinations comparable to that of the pre-exposed subjects. Notably, however that volunteer worked in a high-risk environment during the first months of the pandemic, and asymptomatic infection cannot be ruled out despite the absence of measurable S-specific and neutralizing antibody titers at baseline.

Our study revealed two significant findings regarding vaccination. First is the rapid decline of anti-S antibodies just 40 to 80 days (for unexposed or pre-exposed cohorts, respectively) after a boost with the mRNA vaccine formulations. Second is the sustained level of neutralization ability in the same period that anti-S antibodies are declining. This pattern is the same as the one observed following naturally acquired SARS-CoV-2 infection in 59 subjects. In addition, we observed that—while in both groups the decline of the total anti-S antibodies and IgG titers was significant—the decline in titers was more precipitous in the unexposed group relative to the pre-exposed group. Also highly significant is the observation that the baseline neutralizing activity—but not the total antibody titers—was significantly higher among pre-exposed individuals than the neutralization capacity induced by the first vaccine dose in the unexposed group. This finding is reinforced by the fact that the time after natural infection and the sample use as baseline before the vaccination was more than 4.7 months in average for all 10 pre-exposed subjects. Our results also confirm that antibodies generated after the natural infection, while similar in quantity, are significantly better in their function when natural infection preceded vaccination. These results suggest that natural infection with SARS-CoV-2 may contribute to the expansion of memory B cells, enabling the production of more S-specific antibodies following vaccination. Together, these findings highlight the value of measuring both the function and quantity of S-specific antibodies to follow up humoral immune responses to the vaccination. Our results agree with recent work wherein a predictive model of immune protection from COVID-19 found that the level of neutralizing antibodies is highly predictive of immune protection from symptomatic SARS-CoV-2 infection (Khoury, Cromer, Reynaldi, Schlub, Wheatley, Juno, Subbarao, Kent, Triccas and Davenport 2021) and associated to recovery (Dispinseri, Secchi, Pirillo, Tolazzi, Borghi, Brigatti, De Angelis, Baratella, Bazzigaluppi, Venturi, Sironi, Canitano, Marzinotto, Tresoldi, Ciceri, Piemonti, Negri, Cara, Lampasona and Scarlatti 2021).

Our results on neutralization are built on using the RBD sequence from the original SARS-CoV-2 virus. We do not know the variants infecting the subjects. However, all 59 subjects in the serial sample’s cohort were exposed to the SARS-CoV-2 from March to December 2020. Only the 3 additional subjects in the pre-exposed and vaccinated cohorts were confirmed as positive in the first two weeks of January 2021. During that period information about the circulating variants in Puerto Rico was very limited. The first variant identified in Puerto Rico was the Alpha variant (first identified in the UK, B.1.1.7) and was reported on January 28th, 2021. In addition, from March 2020 to December 2020 the Government of Puerto Rico imposed a strict lockdown limiting the travels to the island requiring mandatory testing upon arrival. By July 21st, 2021, reports from the Surveillance System from the PR Department of Health and other private institutions reported about 950 cases, with patients infected with at least nine (9) different variants as follows: UK Alpha (B.1.1.7), New York (B.1.526), Brazil Gamma (P.1), California Epsilon (B.1.429) and (B.1.427), California Eta (B.1.525), India Delta (B.1.617), Brazil Zeta (P.2), Sudafrica Beta (B.1,351), India Kappa (B.1.617). We acknowledge that the neutralizing properties of our samples may be modified when tested against the RBD from the variant of interest and variant of concerns. However, a work testing four variants representing the original SARS-CoV-2 strain and emerging variants with mutations in the spike protein suggested that infection-and vaccine-induced immunity may be retained against the B.1.1.7 variant (Edara et al. 2021).

Of interest is the role of previous natural infection in driving antibody isotype switching. Particularly in the case of IgA, our results showed that previous exposure led to a faster increase in IgA titers after the first dose of vaccination, while unexposed subjects required a second dose of vaccine to reach same levels of IgA titer of those pre-exposed to the novel coronavirus.

Another critical aspect to be considered is the timing between the natural infection and a potential vaccination against COVID-19. In accordance with the findings of other groups, we highlighted the relevance of the time elapsed between infections or immunizations to induce an optimal immune response (Miller et al. 2008, Pulendran and Ahmed 2006, Serrano-Collazo, Pérez-Guzmán, Pantoja, Hassert, Rodríguez, Giavedoni, Hodara, Parodi, Cruz, Arana, Martínez, White, Brien, de Silva, Pinto and Sariol 2020). Taking into account the results presented here and those from previous works (Bradley, Grundberg, Selvarangan, LeMaster, Fraley, Banerjee, Belden, Louiselle, Nolte, Biswell, Pastinen, Myers and Schuster 2021, Kumar et al. 2020, Prendecki, Clarke, Brown, Cox, Gleeson, Guckian, Randell, Pria, Lightstone, Xu, Barclay, McAdoo, Kelleher and Willicombe 2021), and considering the limited vaccine availability worldwide, our findings suggest that immunity conferred by a single dose may be sufficient to provide immune protection from severe disease in previously-exposed individuals. With this in mind, second doses in previously exposed individuals may be deferred until the final phases of vaccination campaigns and/or to be executed not before than 6 months after the documented infection. Because of the limited number of samples, we were unable to identify any significant differences between the Pfizer-BioNTech or Moderna vaccine formulations.

We are aware of the limitations of this work owing to the limited number of participants and associated clinical data. We also understand that this work would benefit from an examination of the T cell compartment in unexposed and pre-exposed vaccinees, particularly in light of recent evidence that simple serological tests for SARS-CoV-2 antibodies do not reflect the richness and durability of immune memory to SARS-CoV-2 (Dan, Mateus, Kato, Hastie, Yu, Faliti, Grifoni, Ramirez, Haupt, Frazier, Nakao, Rayaprolu, Rawlings, Peters, Krammer, Simon, Saphire, Smith, Weiskopf, Sette and Crotty 2021). With this in mind, experiments characterizing the T cell response in our cohorts are underway.

Nevertheless, this work provides new and additional insight to the limited available data on COVID-19 immune phenomena. Furthermore, this work also advances our understanding of immune responses to the mRNA vaccine formulations in unexposed and pre-exposed individuals, outside of the data provided by the vaccine manufactures. From our results, as well as others (Bradley, Grundberg, Selvarangan, LeMaster, Fraley, Banerjee, Belden, Louiselle, Nolte, Biswell, Pastinen, Myers and Schuster 2021, Khoury, Cromer, Reynaldi, Schlub, Wheatley, Juno, Subbarao, Kent, Triccas and Davenport 2021, Krammer, Srivastava, Alshammary, Amoako, Awawda, Beach, Bermúdez-González, Bielak, Carreño, Chernet, Eaker, Ferreri, Floda, Gleason, Hamburger, Jiang, Kleiner, Jurczyszak, Matthews, Mendez, Nabeel, Mulder, Raskin, Russo, Salimbangon, Saksena, Shin, Singh, Sominsky, Stadlbauer, Wajnberg and Simon 2021, Prendecki, Clarke, Brown, Cox, Gleeson, Guckian, Randell, Pria, Lightstone, Xu, Barclay, McAdoo, Kelleher and Willicombe 2021), the usefulness of a second vaccine dose in pre-exposed subjects remains inconclusive. Furthermore, the immune response elicited by these vaccine formulations needs to be further evaluated to include the T cell compartment, which serves as a critical component in the response to SARS-CoV-2 (Dan, Mateus, Kato, Hastie, Yu, Faliti, Grifoni, Ramirez, Haupt, Frazier, Nakao, Rayaprolu, Rawlings, Peters, Krammer, Simon, Saphire, Smith, Weiskopf, Sette and Crotty 2021, Grifoni et al. 2020, Prendecki, Clarke, Brown, Cox, Gleeson, Guckian, Randell, Pria, Lightstone, Xu, Barclay, McAdoo, Kelleher and Willicombe 2021, Weiskopf et al. 2020). Undoubtably, natural infection confers a strong and high quality humoral and cellular immune response (Dan, Mateus, Kato, Hastie, Yu, Faliti, Grifoni, Ramirez, Haupt, Frazier, Nakao, Rayaprolu, Rawlings, Peters, Krammer, Simon, Saphire, Smith, Weiskopf, Sette and Crotty 2021, Goldberg et al. 2021, Grifoni, Weiskopf, Ramirez, Mateus, Dan, Moderbacher, Rawlings, Sutherland, Premkumar, Jadi, Marrama, de Silva, Frazier, Carlin, Greenbaum, Peters, Krammer, Smith, Crotty and Sette 2020). This fact has recently been underscored by work showing that variants of concern partially escape humoral—but not T-cell-mediated—immune responses in COVID-19 convalescent donors and vaccinees (Geers, Shamier, Bogers, den Hartog, Gommers, Nieuwkoop, Schmitz, Rijsbergen, van Osch, Dijkhuizen, Smits, Comvalius, van Mourik, Caniels, van Gils, Sanders, Oude Munnink, Molenkamp, de Jager, Haagmans, de Swart, Koopmans, van Binnendijk, de Vries and GeurtsvanKessel 2021). As the CDC’s guidelines on the impact of the vaccination on our lifestyles (travel quarantine and testing, maskless outside and indoors) continues to change and evolve, it is remains unclear why immunity conferred by natural infection is not taken in to account to support those guidelines, nor it is considered in the progress towards attaining herd-immunity that may enable us to return to the new social normality. In this context, our results are also highly relevant to consider standardizing methods that both serve as a tool to follow up the immune response to the vaccination, but also to provide a correlate of protection.

## Data Availability

All additional data collected during this study is available as supplementary information

## Authors Contribution

CAS and AME conceptualized the work and supervised the studies. PP supervised the work and performed the serologic, neutralization test and supported the figures design. CSC, TRA, AA execute the serological work. CC and GL selected samples from blood donors. JDB, AKP, CC, GL, PP contribute to the results discussion and analysis. DA, CPC, PP coordinate and supervise the cohort’s management and follow up. PP and TRA organized the data for future analysis. TA provided administrative and regulatory support. JDB and AKP designed and supervised the BSL3 work. ETS performed FRNT analysis. CAS wrote the initial draft, with the other authors providing insights and concepts. ETS and AKP conducted the editorial work for the final manuscript.

## Acknowledgements

Authors want to thank the volunteers that were willing to participate and to contribute to science. To Ilia Toledo, MT, Francheska Rivera, MT and Drs. Ivelisse Martin for their contribution and diligent efforts to provide access to the samples from some presumptive-positive subjects exposed to SARS-CoV-2. Particular acknowledgement is deserved for all administrative and supportive staff at the Medical Sciences Campus, University of Puerto Rico, Laboratorio Clinico Toledo, Laboratorio, Clinico Martin, Banco de Sangre Centro Médico and Banco de Sangre Servicios Mutuos for their availability and commitment during the curfew imposed by the quarantine period. Thanks also to the Latin clinical Trial Center’ staff for their dedication providing excellent care to the participants.

The Puerto Rico Science, Technology and Research Trust supported research reported in this work under agreement number 2020-00272 to AME and CAS. Also, the University of Puerto Rico contributed with the UPR-COVID-19 Grant to CAS and AME. This work was also supported by 1U01CA260541-01 to CAS (NCI/NIAID). This work was also partiallly funded by Saint Louis University COVID-19 research Seed Funding to awarded to AKP and awarded to JDB.

## FIGURE LEGENDS

**Supplementary Figure S1: Antibody subclasses isotypes in a longitudinal cohort of 59 volunteers exposed to SARS-CoV-2.** Panel A shows the total anti-S antibodies in the second set of samples collected an average of 67.8 days after the first set of samples (an average of 108 days after PCR-confirmed SARS-CoV-2 infection). A third sample was collected from a subset of the participants (n=12) an average of 99.5 days after the second set of samples (an average of 207 days after infection). Two different patterns in the kinetics of the antibody response were identified: (1) 74.5% of samples showed a decrease in the binding from the time of the first to the second sampling (Panel B) and (2) 25.4% of samples showed increased values relative to the first sampling (Panel C). Panels D-G show the results of antibody binding for the different subclasses tested, with IgG1 being the predominant subclass. Panels H and I show the results for IgM and IgA isotypes. Statistical significance was determined by two-way ANOVA multiple comparisons to test for increase or decrease among samples. p<0.05 was considered significant. Sample 3 encompass the 15 subjects from whom a collection of a third sample was completed. Panels D to I, includes the number of samples, from the initial cohort of 59 subjects before vaccination, that were positive for each of the antibody’s subtype or subclasses as described in the results section.

**Supplementary Figure S2: Time elapsed between diagnosis and sample collection was not significantly different between groups.** There were no significant differences in the time from diagnostic (Dx) to the first sample collection or between the first and the second samples collection in both groups. Statistical significance was determined by two-way ANOVA multiple comparisons was used. p<0.05 was considered significant. Results are from the 59 subjects in the initial cohort before vaccination. From two subjects in the increased titer and from one in the decreased subgroups we were unable to establish the precise time of diagnostic.

**Supplementary Figure S3: IgG titers—but not IgM or IgA—correlate with neutralization.** Panel A shows the correlation between the neutralization capacity measured with the surrogate viral neutralization test (sVNT) and the total IgG titers, confirming a moderate agreement. Panel B also shows moderate agreement between the sVNT and Focus Reduction Neutralization Tests (FRNT) using the whole virus. Panels C and D show a fair and a slight agreement between the neutralization activity and the IgM and IgA titers, respectively. All samples (n=131) from the 59 subjects in the initial cohort, before vaccination, were included in the analysis for figures in panels A, C and D. A subset of 15 samples with prior known FRNT results, were used for the correlation analysis showed in panel B. Cohen’s Kappa agreement follow Landis and Koch scale. The values (κ) were considered as follows: poor agreement, κ<0.02); fair agreement, κ=0.21 to 0.4; moderate agreement, κ=0.41 to 0.6; substantial agreement, κ=0.61 to 0.8; very good agreement, κ=0.81 to 1.0

**Supplementary Figure S4: Time elapsed between sample collection after vaccination.** The time between the first and second samples after the 1^st^ or the 2^nd^ vaccine dose (1S-P1st-vd, 1S-P2st-vd) were similar in both groups (pre-exposed and unexposed vaccinated subgroups). However, the time of collection of the third sample (2S-P2nd-vd) was significantly longer for the pre-exposed group compared with the unexposed group. Statistical significance was determined by two-way ANOVA multiple comparisons were used. *p<*0.05 was considered significant. Unexposed and vaccinated group n=21. Pre-exposed and vaccinated group n=10.

**Supplementary Figure S5: IgG1, IgM and IgA are differentially boosted by the vaccination in healthy or pre-exposed vaccinated subgroups.** The boost of the IgG1 in both subgroups agrees with the total antibodies’ changes showed in figure 3 after each vaccine dose. First vaccine dose induces a significant increase in the IgM values only in the unexposed healthy subjects. The first vaccine dose significantly boosted the IgA values in both groups. The increase in IgA titers was significantly higher in the pre-exposed vaccinated group compared to the healthy-vaccinated group. The second vaccine boost resulted in an additional significant increase in IgA titers only in the healthy-vaccinated group suggesting an advantage of the second shot in naïve individuals.

