## Supplementary Figures for "Function is more reliable than quantity to follow up the humoral response to the Receptor Binding Domain of SARS-CoV-2 Spike protein after natural infection or COVID-19 vaccination"

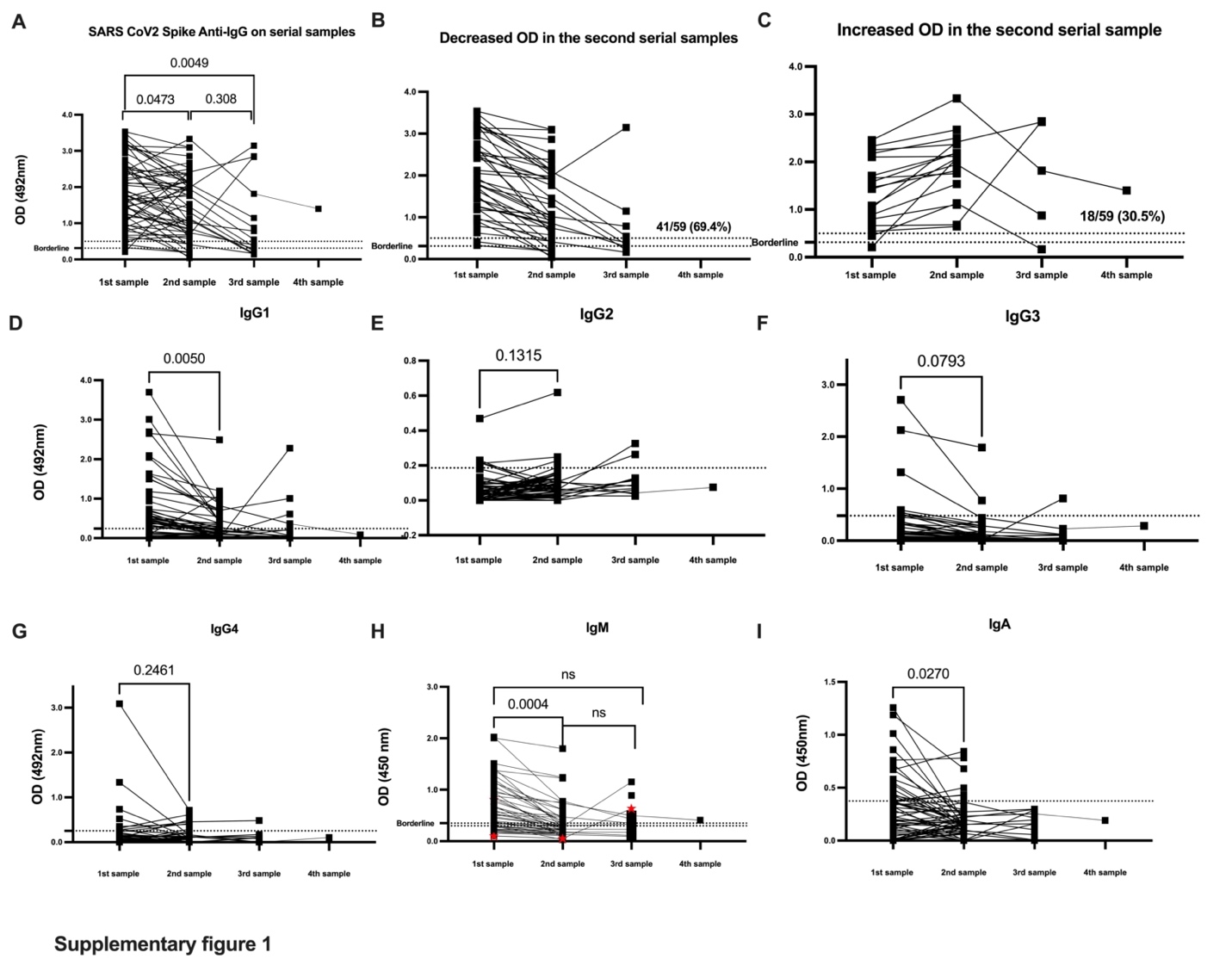

**Supplementary Figure S1: Antibody subclasses isotypes in a longitudinal cohort of 59 volunteers exposed to SARS-CoV-2.** Panel A shows the total anti-S antibodies in the second set of samples collected an average of 67.8 days after the first set of samples (an average of 108 days after PCR-confirmed SARS-CoV-2 infection). A third sample was collected from a subset of the participants (n=12) an average of 99.5 days after the second set of samples (an average of 207 days after infection). Two different patterns in the kinetics of the antibody response were identified: (1) 74.5% of samples showed a decrease in the binding from the time of the first to the second sampling (Panel B) and (2) 25.4% of samples showed increased values relative to the first sampling (Panel C). Panels D-G show the results of antibody binding for the different subclasses tested, with IgG1 being the predominant subclass. Panels H and I show the results for IgM and IgA isotypes. Statistical significance was determined by two-way ANOVA multiple comparisons to test for increase or decrease among samples. p<0.05 was considered significant. In Panel A, samples 1 and 2 include the 59 subjects in the initial cohort before vaccination. Sample 3 encompass the 15 subjects from whom a collection of a third sample was completed. Panels D to I, includes the number of samples, from the initial cohort of 59 subjects before vaccination, that were positive for each of the antibodies subtype or subclasses as described in the results section.

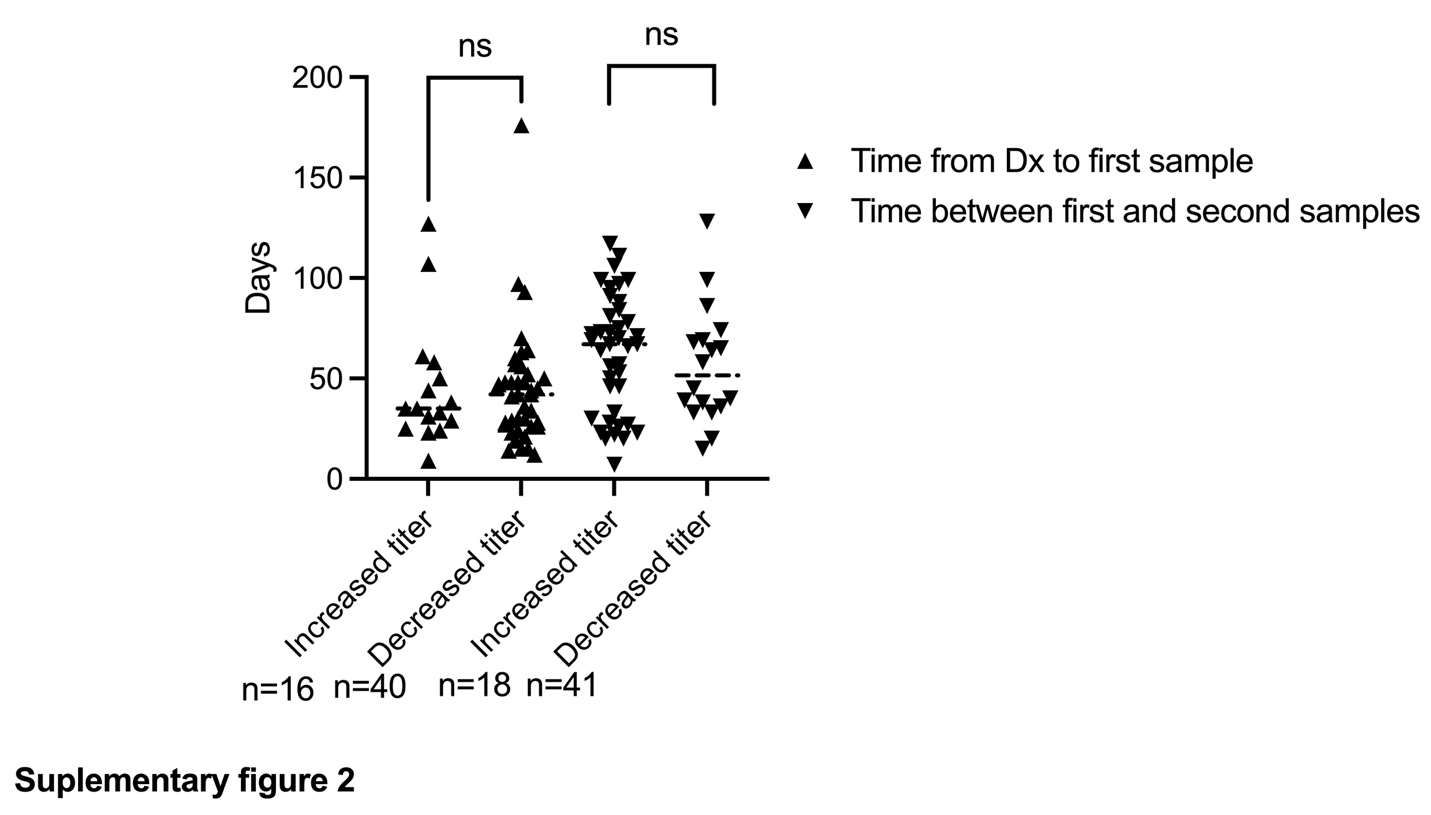

**Supplementary Figure S2: Time elapsed between diagnosis and sample collection was not significantly different between groups.** There were no significant differences in the time from diagnostic (Dx) to the first sample collection or between the first and the second samples collection in both groups. Statistical significance was determined by two-way ANOVA multiple comparisons was used. p<0.05 was considered significant. Results are from the 59 subjects in the initial cohort before vaccination. From two subjects in the increased titer and from one in the decreased subgroups we were unable to establish the precise time of diagnostic.

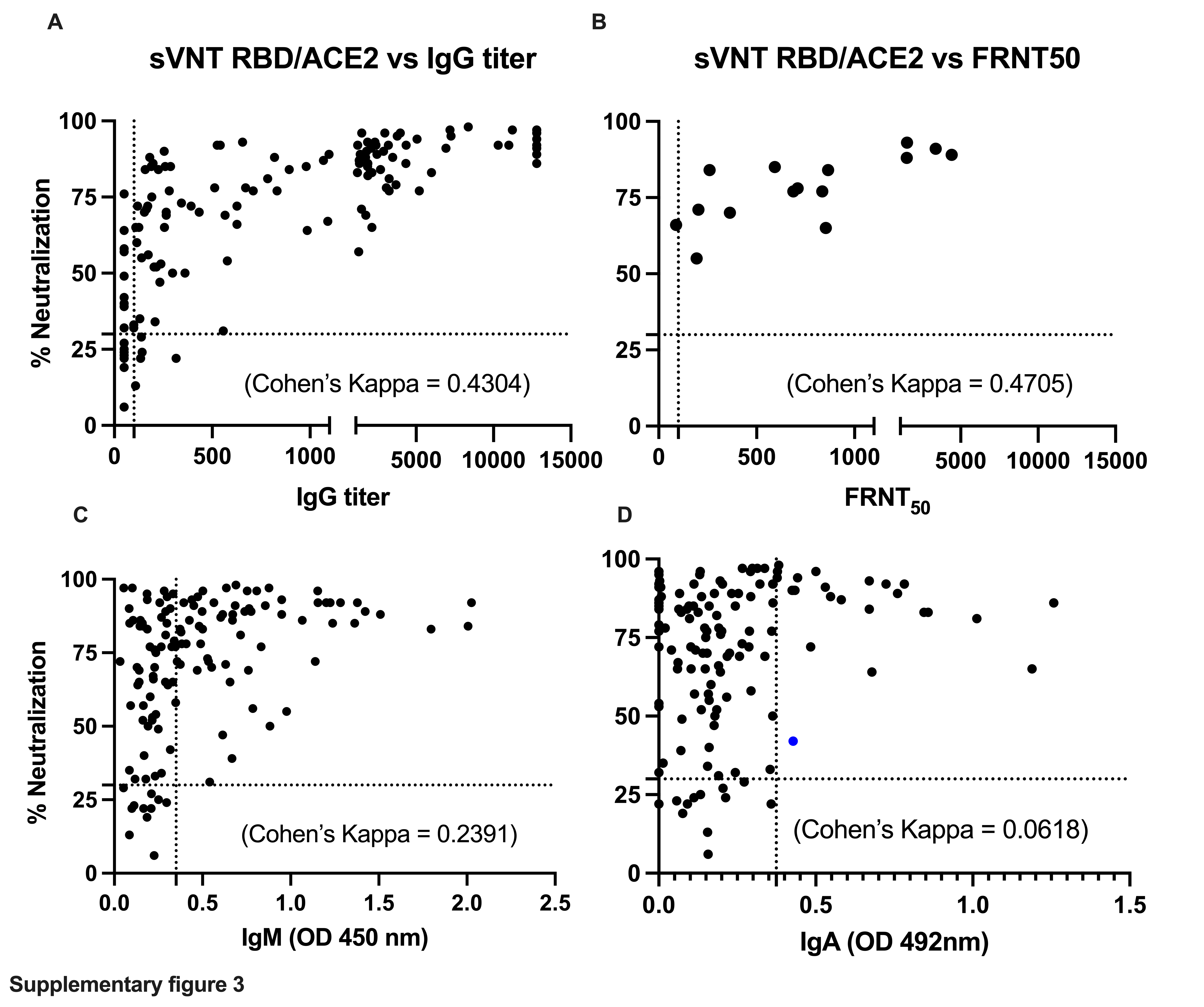

**Supplementary Figure S3: IgG titers—but not IgM or IgA—correlate with neutralization.** Panel A shows the correlation between the neutralization capacity measured with the surrogate viral neutralization test (sVNT) and the total IgG titers, confirming a moderate agreement. Panel B also shows moderate agreement between the sVNT and Focus Reduction Neutralization Tests (FRNT) using the whole virus. Panels C and D show a fair and a slight agreement between the neutralization activity and the IgM and IgA titers, respectively. All samples (n=131) from the 59 subjects in the initial cohort, before vaccination, were included in the analysis for figures in panels A, C and D. A subset of 15 samples with prior known FRNT results, were used for the correlation analysis showed in panel B.

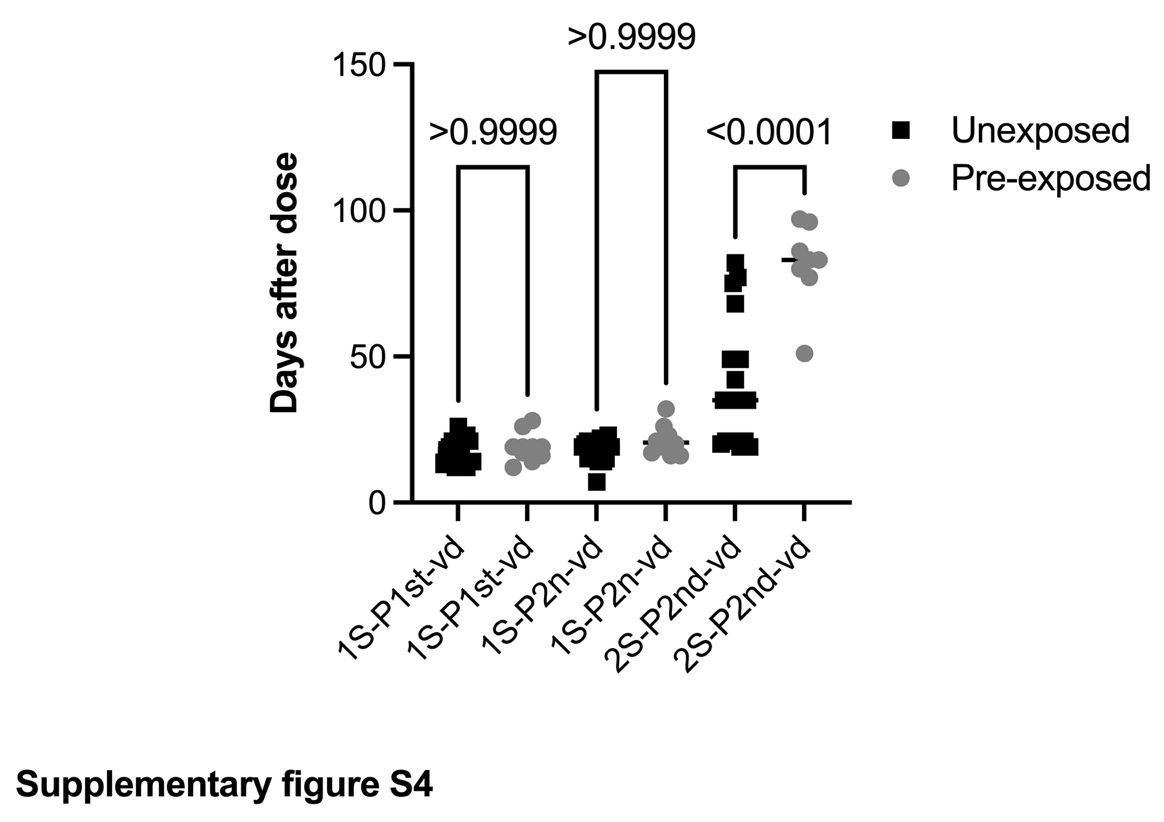

**Supplementary Figure S4: Time elapsed between sample collection after vaccination.** The time between the first and second samples after the 1^st^ or the 2^nd^ vaccine dose (1S-P1st-vd, 1S-P2st-vd) were similar in both groups (pre-exposed and unexposed vaccinated subgroups). However, the time of collection of the third sample (2S-P2nd-vd) was significantly longer for the pre-exposed group compared with the unexposed group. Statistical significance was determined by two-way ANOVA multiple comparisons were used. *p<*0.05 was considered significant. Unexposed and vaccinated group n=21. Pre-exposed and vaccinated group n=10.

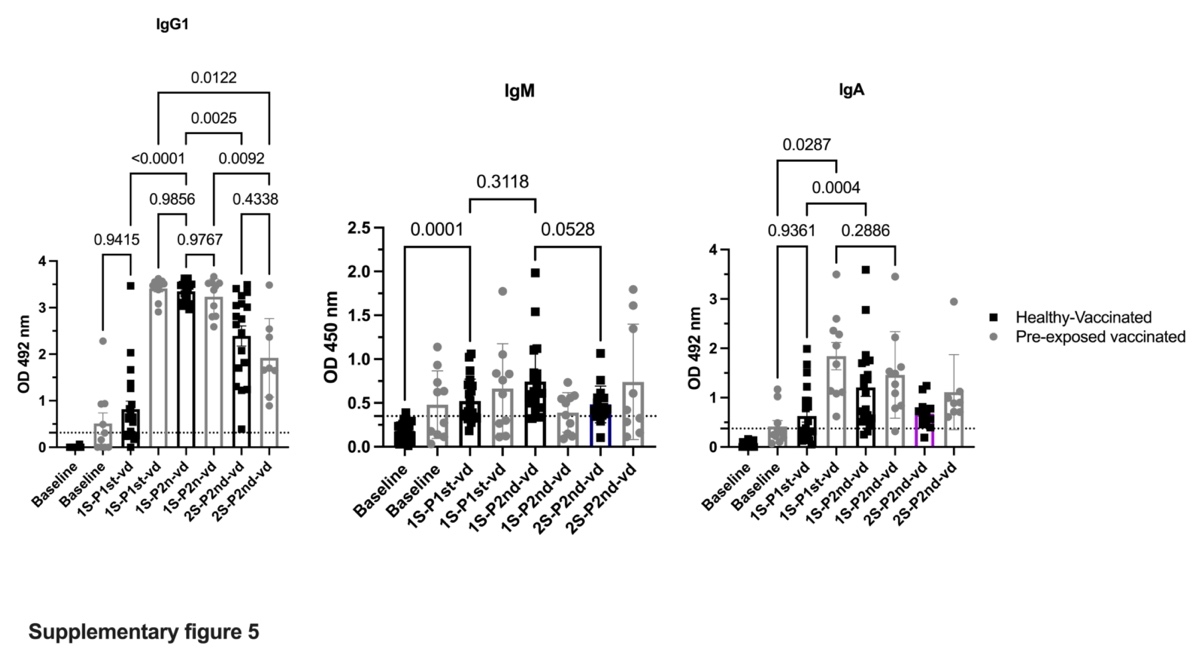

**Supplementary Figure S5: IgG1, IgM and IgA are differentially boosted by the vaccination in healthy or pre-exposed vaccinated subgroups.** The boost of the IgG1 in both subgroups agrees with the total antibodies’ changes showed in figure 3 after each vaccine dose. First vaccine dose induces a significant increase in the IgM values only in the unexposed healthy subjects. The first vaccine dose significantly boosted the IgA values in both groups. The increase in IgA titers was significantly higher in the pre-exposed vaccinated group compared to the healthy-vaccinated group. The second vaccine boost resulted in an additional significant increase in IgA titers only in the healthy-vaccinated group suggesting an advantage of the second shot in naïve individuals.
